# Effectiveness of three-year old piperonyl butoxide and pyrethroid-treated long-lasting insecticidal nets (LLINs) versus pyrethroid-only LLINs against malaria infection: results of a cluster randomised trial in Tanzania

**DOI:** 10.1101/2022.07.06.22277292

**Authors:** Natacha Protopopoff, Jacklin F. Mosha, Louisa A. Messenger, Eliud Lukole, Jacques D. Charlwood, Alexandra Wright, Enock Kessy, Alphaxard Manjurano, Franklin W. Mosha, Immo Kleinschmidt, Mark Rowland

**Affiliations:** Department of Disease Control, London School of Hygiene and Tropical Medicine, London, United Kingdom; National Institute for Medical Research, Mwanza Medical Research Centre, Mwanza, Tanzania; Kilimanjaro Christian Medical University College, Moshi, Tanzania; MRC International Statistics and Epidemiology Group, London School of Hygiene and Tropical Medicine, London, United Kingdom; School of Pathology, Faculty of Health Sciences, University of Witwatersrand, South Africa

## Abstract

**Background:** After decades of success in reducing malaria through the scale-up of pyrethroid long-lasting insecticidal nets (LLINs), malaria decline has stalled, coinciding with the rapid spread of pyrethroid resistance. A new class of net, treated with a mixture of a pyrethroid and a synergist, piperonyl butoxide (PBO), demonstrated superior efficacy compared to standard (std) pyrethroid LLINs against malaria in an area of intense pyrethroid resistance, reducing malaria prevalence by 44% over 2 years in the present trial. However, an important question is left unanswered regarding the performance of this PBO-LLIN over the World Health Organization recommended lifespan of 3 years for LLINs.

**Methods and Findings:** We conducted a four-arm randomized controlled trial using a two-by-two factorial design that evaluated the effectiveness of PBO-LLIN arms (12 clusters PBO-LLIN alone and 12 clusters PBO-LLIN + Indoor Residual Spraying; IRS) compared to std-LLIN (12 clusters std-LLIN alone and 12 clusters std-LLIN + IRS) and IRS arms versus no IRS arms from January 2014 to December 2017 in Muleba, Tanzania. Malaria infection prevalence in 80 children, 6 months to 14 years, per cluster was measured twice a year and analysed in an intention to treat (ITT) and per protocol (PP) approach. Density of malaria mosquito vectors and entomological inoculation rate (EIR) were assessed monthly in 7 houses per cluster. Logistic regression allowing for within cluster correlation of responses was used to compare malaria prevalence between PBO-LLIN groups vs std-LLIN groups and IRS groups vs no IRS groups during the third-year follow-up at 28- and 33-months post-intervention. No further IRS was conducted after the first spray round in 2015; as yearly IRS is recommended by WHO, results need to be interpreted in light of this limitation. Vector density and EIR were analysed using negative binomial regression. Malaria results were available for 7471 children. At 28 months, malaria infection prevalence was lower in the PBO-LLIN groups (69.3%) compared to the std-LLIN groups (80.9%, Odds Ratio: 0.45, 95% Confidence Interval: 0.21-0.95, p value: 0.0364). The effect was weaker at 33 months post-intervention (OR: 0.60, 95%CI:0.32-1.13, p value: 0.1131), in the ITT analysis but still evident in the PP analysis (OR: 0.34, 95%CI: 0.16-0.71, p value: 0.0051). At this time point, net usage in household participants was 31% and PBO concentration in PBO-LLINs was reduced by 96% compared to those of new nets. A total of 17,451 *Anopheles* mosquitoes were collected during the 3150 collection nights done in the third year. There was no reduction in EIR (DR: 0.63, 95%CI: 0.25-1.61, p value: 0.3296) between the PBO groups and std-LLIN groups or between IRS and no IRS groups (DR: 0.7, 95%CI: 0.41-2.28, p value: 0.9426).

**Conclusions:** PBO-LLINs no longer provided community protection from malaria infection, compared to std-LLINs by the third year of use due to low net usage. Children still sleeping under PBO-LLINs had lower odds of infection than those sleeping under a std-LLIN, however prevalence remained unacceptably high. It is urgent that net distribution frequencies and effective lifespan of this class of LLIN are aligned for maximum impact.

**Trial registration:** ClinicalTrials.gov NCT02288637

**Author summary:** *Why was the study done?:*

- Widespread insecticide resistance among major malaria mosquito populations threatens control efforts worldwide.
- A new class of long-lasting insecticidal net (LLIN), containing a pyrethroid insecticide and a synergist, piperonyl butoxide (PBO), improves insecticide toxicity by inhibiting metabolic enzyme activity, responsible for insecticide resistance.
- PBO-LLINs reduced malaria prevalence by 44% in Tanzania and 27% in Uganda, compared to standard pyrethroid-only LLINs, in two 24-month cluster randomised controlled trials (CRTs), as conducted per World Health Organization (WHO) recommendations.
- However, LLIN deployment regimens are currently based on 3 years assumed functional survival for nets, with questions remaining, regarding the effectiveness of PBO-LLINs to prevent malaria after 3 years of continuous field use.

*What did the researchers do and find?:*

- A 24-month CRT in Muleba, Tanzania, an area of high pyrethroid resistance, was extended for one year to assess the impact of PBO-LLINs, compared to pyrethroid-only LLINs, on malaria infection, after 3 years of use, corresponding to their expected lifespan.
- After 28 months, malaria infection prevalence was still lower in users of PBO-LLINs, compared to standard LLINs, but this effect was lost by 33 months in the intention to treat (ITT) analysis but was still evident in the per protocol (PP) analysis.
- No reduction in vector density or entomological inoculation rate was evident after 3 years of use.
- Reasons underlying the poorer PBO-LLIN performance after 36 months, included declining net usage, poor netting durability and diminished PBO synergist content.

*What do these findings mean?:*

- By 3 years of continuous field use, PBO-LLINs no longer provided community protection from malaria infection, compared to pyrethroid-only LLINs, however children sleeping under a PBO-LLIN still retained a level of protection compared to those sleeping under a standard pyrethroid-only LLIN.
- To maximise the impact of PBO-LLINs, net procurement and replacement strategies, must be modified to maintain high coverage.
- Future CRTs and additional community studies are required to determine the effective lifespan of new classes of LLINs for appropriate incorporation into resistance management schemes, to preserve gains made in malaria control over the past two decades.

## Introduction

Globally, significant progress has been made controlling malaria through the scale-up of long-lasting insecticidal nets (LLINs), indoor residual spraying (IRS) and other prevention, diagnostic and treatment tools [1]. In the past two decades, the World Health Organization (WHO) estimated that 1.5 billion malaria cases and 7.6 million malaria deaths were averted due to the implementation of large scale interventions, of which 68% has been attributed to LLINs [2]. In recent years the decline in malaria cases has started to reverse in many countries due to a number of factors. Insecticide resistance, particularly to pyrethroid insecticides, has become widespread, thereby undermining the effectiveness of standard LLINs to control malaria [2]. In response, substantial investments have been made in the development of new insecticides and chemical classes, with novel modes of action, in an effort to improve malaria vector control and to potentially mitigate further selection for insecticide resistance [3].

Until 2017, pyrethroids were the only chemical recommend by the WHO for use in LLINs. The first new class of dual-active-ingredient LLINs were nets that were treated with a mixture of a pyrethroid insecticide and a synergist, piperonyl butoxide (PBO), which enhances insecticide toxicity by inhibiting the activity of metabolic enzymes, commonly over-expressed in resistant vector populations [4, 5]. PBO-LLINs received a WHO recommendation following successful evaluation in two cluster randomised controlled trials (CRTs), after 24 months of community-use [6]. In these initial CRTs, PBO-LLINs reduced community malaria infection prevalence by 44% in Tanzania [7] and 27% in Uganda [8], compared to standard pyrethroid-only LLINs in areas of high pyrethroid resistance. A subsequent meta-analysis determined that PBO-LLINs reduced the odds of malaria infection by 31% compared to standard pyrethroid-only LLINs 21-25 months after deployment [9]. PBO-LLINs are now being distributed by National Malaria Control Programmes (NMCPs) across sub-Saharan Africa [10], with just 7,000 nets procured in 2016 to over 2 million in 2019 [11]. Concurrent rollouts by the President’s Malaria Initiative estimated that in the fiscal year 2020, 30% of their deployed LLINs in Africa contained PBO and this figure is set to rise. A second generation of LLINs combining a pyrethroid and a pyrrole (chlorfenapyr) or insect growth regulator (pyriproxyfen) have also undergone CRT evaluations [12, 13], with the former LLIN reducing malaria infection prevalence by 55% after 2 years in Tanzania [12].

Net deployment regimens currently assume three years of functional survival for LLINs under field conditions [14]. However, multiple cohort studies of pyrethroid LLINs across Africa have demonstrated substantial heterogeneity in effective net lifespan, both within and between types of LLIN and endemic regions; net surveys from 13 countries estimated a median functional net survivorship of 40%, three years after distribution [15]. These observations reflect variations in hole accumulation and rates of insecticide decline, which while initially dictated by product textile and chemical features, are in turn exacerbated by differences in user behaviour and acceptability [16-18]. The deterioration characteristics of pyrethroid-only LLINs cannot be extrapolated to dual-active-ingredient LLINs, because partner synergists or insecticides may degrade faster than pyrethroids. A household randomised trial from Kenya reported more than 80% of PBO was lost from PBO-LLINs after 3 years of use compared to 50% of the pyrethroid insecticide [19].

Overall, there is a paucity of field data evaluating the efficacy of aged PBO-LLINs to prevent malaria. Their superiority and potential for cost-effectiveness over pyrethroid-only LLINs [20], while evident after 2 years in some endemic settings, may decline rapidly thereafter. In a recent CRT conducted in a different region of Tanzania, the reduction in malaria prevalence between PBO-LLIN compared to standard LLIN was 45% after 1 year, while there was no further reduction observed in the second year [12]. There is clearly a need for large-scale longitudinal studies of PBO-LLINs across disparate communities. Crucial questions remain unanswered regarding the performance of PBO-LLINs after 24 months, which will have direct implications for their frequency of procurement and their role in evidence-based resistance management schemes, in the current arsenal of dual-active-ingredient LLINs and IRS products. This paper reports the results of a trial of the effectiveness of PBO-LLINs compared to pyrethroid-only LLINs against malaria infection after 3 years of continuous community-use in an area of intense pyrethroid resistance in northern Tanzania.

## Methods

Details of the study and its main outcomes relating to two years of follow-up have been reported previously [7]. The main study concluded in 2016 but follow-up was extended for an additional year to assess the effectiveness of PBO-LLINs during a third year of use in the community which corresponds to their expected full lifespan. There were no further study interventions during this period although study participants in all trial arms received standard of care LLIN top-ups through antenatal care clinics and school distributions, as and when they were eligible. Ethical approval was obtained from the Kilimanjaro Christian Medical University College (registration 781), the London School of Hygiene and Tropical Medicine (reference 6551), and the Tanzanian National Institute for Medical Research (registration NIMR/HQ/R.8a/VolIX/1803). Written informed consent was provided by an adult caregiver of participating children. The trial was registered with ClinicalTrials.gov (registration number NCT02288637). Consolidated Standards of Reporting Trials (CONSORT) checklist are available as supporting documents (Table S1).

### Study design

We conducted a four-arm CRT using a two-by-two factorial design between January 2014 and December 2017 in 40 villages in Muleba district, Kagera region, northern Tanzania [7]. Clusters, divided into an inner core area, and an outer buffer zone of 300 metres minimum, were randomized in a 1:1:1:1 ratio to the following treatments: standard LLIN; PBO-LLIN; standard LLIN + IRS and PBO-LLIN + IRS.

### Participants and data collections

All households in the study area were eligible to receive the interventions. Only households situated in the cluster core area with children aged 6 months to 14 years were eligible for inclusion in malaria and mosquito cross-sectional surveys conducted twice a year (June and November) between 2015 and 2017. Exclusion criteria included dwellings not found or vacant, unwillingness to give informed consent, or eligible children who were severely ill or who did not reside permanently in the household.

During each cross-sectional survey 55 households meeting the eligibility requirement were randomly sampled in each cluster. Household characteristics and information about number of residents, household wealth and insecticide-treated net (ITN) ownership and usage was collected using an electronic form programmed to run on personal digital assistants (PDAs) with Pendragon software (Universal version, Pendragon Software Corporation, Chicago, United States). In each household, up to three eligible children were selected at random for malaria *Plasmodium falciparum* infection testing using rapid diagnostic tests (CareStart™ (pf/PAN) Combo Test, DiaSys, Workingham, UK). Children found positive for malaria were treated with artemether–lumefantrine according to national guidelines. Haemoglobin concentration was also measured (HemoCue(R) Hb 201+ Ängelholm, Sweden) to assess anaemia prevalence. Children were checked for other symptoms and treated accordingly or referred to health facilities. The first (2015) and second year (2016) follow-up surveys have been reported elsewhere [7]. This paper reports data from surveys conducted at 28 months (June 2017) and 33 months (November 2017) post LLIN distribution.

Monthly entomological cross-sectional surveys were conducted during the 3 years post intervention, with collections carried out between 03 January 2017 and 15 December 2017 for the third year. For each collection round, seven households per cluster were selected at random and a Center for Disease Control and Prevention (CDC) light trap was installed at the foot of a bed in one of the bedrooms and monitored for one night to assess vector density. Existing nets were not removed, but a new standard LLIN was hung if the bed selected for CDC light trap had no net. Anophelines were morphologically identified to species or species group [21] and PCR Taq Man assay [23] was used to distinguish the two sibling species *An. gambiae* and *An. arabiensis*. A subset of malaria vectors was tested for *P. falciparum* circumsporozoite protein (CSP) to estimate sporozoite prevalence and calculate entomological inoculation rate (EIR) [22].

### Interventions

Both IRS and LLIN distributions were conducted in February 2015. 45,000 standard LLIN, Olyset LLIN (Sumitomo Chemical, Japan), containing 20 g/kg of the pyrethroid permethrin, and 45,000 PBO-LLIN, Olyset Plus (Sumitomo Chemical, Japan), combining PBO (10 g/kg) and the pyrethroid permethrin (20 g/kg) were distributed in the allocated study arms to provide one LLIN per two persons. The organophosphate insecticide Actellic® 300CS (Syngenta, Switzerland) containing microencapsulated pirimiphos-methyl, was sprayed onto the interior walls and ceiling or roof of each house at the recommended dosage of 1 g/m^2^, in the two IRS study arms. No further IRS was conducted after the February 2015 spray round. Duration of efficacy of Actellic® 300CS against wild mosquitoes, reported in experimental hut trials done in several countries, ranged between 3 to 9 months [24]. IRS implemented, as per WHO specification, every year would have been necessary for a valid comparison against standard LLIN whose WHO recommended lifespan is 3 years. While the effect of IRS is reported as per analysis plan, interpretation can only be drawn in regards to the implementation and not the quality and residual efficacy of the insecticide.

Insecticide and PBO concentration was assessed in a subset of LLINs, collected after 36 months of use in the community, by high performance liquid chromatography (HPLC) at the Liverpool School of Tropical Medicine [25].

### Randomisation and masking

The 48 clusters were randomised to the four-study arms by an independent epidemiologist and balanced on the following three restriction variables recorded in the baseline survey: malaria infection prevalence in children aged 6 months to 14 years (maximum difference allowed between study arms +/- 7%), ITN usage (+/-10%) and household in the lowest socio-economic status tercile (+/-10%). Of 200,000 random allocations 29,478 met the restriction criteria, and after verifying that clusters were independently allocated to study arms, one of the eligible allocations was randomly chosen [7].

The two LLINs were of same colour and shape and only distinguishable by label codes and different coloured thread stitch during manufacturing process. Participants and field staff collecting the data were blinded to the type of LLINs received but could not be blinded to the IRS treatment allocation.

### Outcomes

The primary outcome was malaria infection prevalence in children 6 month to 14 years of age measured at 28- and 33-months post LLIN distribution. A secondary clinical outcome was the prevalence of severe anaemia (defined as haemoglobin <8 g/dl) in children aged 6 months to 4 years old at the same time points. The main entomological outcome was EIR, during the third year of follow-up, and other endpoints included vector population density and sporozoite prevalence assessed during the same period.

### Sample size and statistical analysis

24 clusters of 80 individuals per cluster provided 80% power to detect a 28% relative reduction in malaria infection prevalence between IRS versus no IRS, and PBO-LLIN versus no PBO-LLIN assuming a mean prevalence of 20% in the reference arms, and a coefficient of variation of 0.3 [7]. This sample size also allowed us to detect a 40% difference between each of the four individual arms (12 clusters per arm).

Differences in malaria infection prevalence and anaemia prevalence (restricted to children from 6 months to 4 years old) between IRS versus no IRS, and PBO-LLIN versus no PBO-LLIN was conducted in an intention to treat (ITT) basis at each time point (28 and 33 months), separately using logistic regression allowing for within cluster correlation of responses by using a robust variance estimator to calculate standard errors. Interaction between IRS and PBO-LLIN was assessed. No allowance was made for multiplicity of testing in the analyses. Per-protocol (PP) analysis, restricted to children using the allocated LLINs, was also conducted for both outcomes.For the entomological outcomes, vector density and EIR, negative binomial regression with a robust variance estimator was used to estimate density rate ratios between groups after adjusting for baseline. EIR was estimated as the mean number of sporozoite infected *Anopheles* per house per night [26] and weighted to account for the proportion of collected *Anopheles* processed for sporozoites. Sporozoite rate was compared using logistic regression.

To assess reduction in malaria prevalence over the 3 years, PBO-LLIN arms (with and without prior IRS) and the same for standard pyrethroid LLIN were combined. A post hoc intention to treat analysis was done using a logistic regression with a robust variance estimator. The model included fixed effects for time, study arm, IRS and the interaction time by study arm. All 6 malaria infection prevalence surveys conducted each year in June and November between 2015-2017 were included in the analysis.

## Results

### Participants

All 29,365 households received the allocated interventions (Figure 1) and 13,568 were eligible to be enrolled for data collection; other households were located in the buffer area or had missing information about their geolocations and were excluded. During each cross-sectional survey, 2640 households were selected, of which 68% (N=1798) gave consent at t28 (14 June-11 July 2017) and 64% (N=1680) at t33 (10 nov-13 December 2017). The most common reasons not to participate was household members not being at home during the visit, with 14% (N=365) of the responses at t28 and 16% (N=435) at t33, households with no children of eligible age (t28=271 and t33=271) and households not found (t28=155 and t33=182). Only 37 at the t28 survey and 40 at the t33 survey of the households refused to participate. 4357 children were enrolled at t28, 89% (N=3884) were tested for malaria prevalence, while the remaining children did not attend testing. At t33, 4117 were enrolled and 87% (N=3594) were tested. Of the children tested in both cross-sectional surveys, 7471 were included in the analysis as 7 had missing malaria results. For mosquito collections, 5184 households were selected at random, of these 3150 (61%) gave consent; 1861 (36%) were not visited as the sample size of 8 households consented per cluster per round was met. The remaining households were either not found, nobody was home during the visit, or refused to participate.

**Figure 2:**
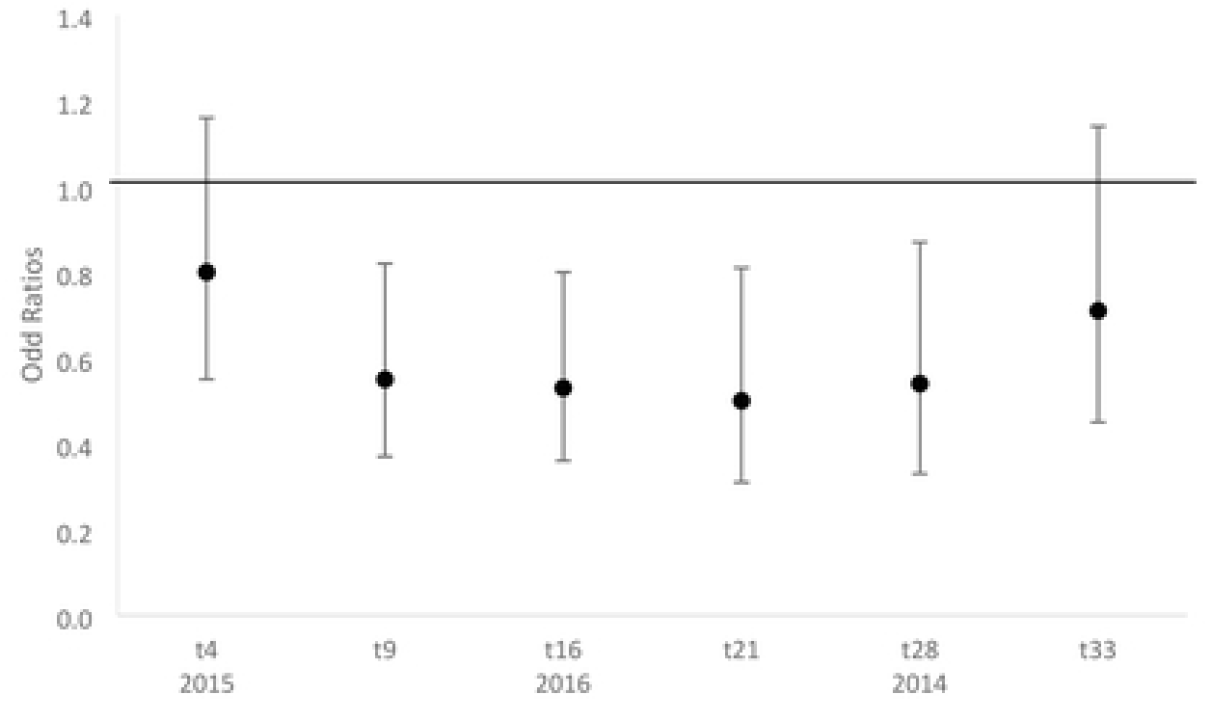
Odd Ratios with 95% confidence interval for malaria prevalence in each cross sectional surveys between PBO LLINs arms (with and without IRS) vs std LLINs arms (with and without IRS). An intention to treat analysis was done using a logistic regression with a robust variance estimator comparing PBO-LLIN arms combined vs std LLIN arms combined and included fixed effects for time, study arm, IRS and the interaction time by study arm. P-value for interaction term was 0.0060.

### Baseline data

Baseline results were presented previously [7], showing that children, household and cluster characteristics were similar between study arms. Malaria prevalence in children aged 6 months to 14 years ranged from 61% to 68% which was within the limits set for the randomisation restriction criteria. Overall usage of study nets in the four groups dropped from 73% (3132/4312) 4 months post distribution to 47% (1847/3946) after 28 months, and further reduced to 31% (3031/9732) after 33 months with minor variation between study arms (highest usage: 34% for standard LLIN group vs lowest: 30% for the PBO-LLIN + IRS arm) (Table S2, Figure S1). Study net usage in enrolled children was similar to that observed in other household members. Usage of any LLIN, which included study nets and standard LLINs received from routine distributions to children through primary school and to pregnant women during antenatal clinic visits, remained around 60% from 16 months onward. After 36 months of use, the concentration of permethrin remaining in tested PBO-LLINs was 9 g/kg compared to 20 g/kg when new, while 10 g/kg of permethrin remained in standard LLINs compared to 20 g/kg when new. PBO was depleted, with 0.4 g/kg of 10 g/kg remaining in the net fibres [25]

### Epidemiological results

At 28 months post-intervention, malaria infection prevalence in the PBO-LLIN arms was lower compared to the standard LLIN arms (80.9%, versus 69.3%, Odds Ratio: 0.45, 95%Confidence Interval: 0.21-0.95, p: 0.0364) in the ITT factorial analysis (Table 1). The effect was no longer significant at 33 months post distribution (OR: 0.60, 95%CI (0.32-1.13), p value: 0.1131). Regarding children who slept under the allocated study nets in the PP analysis, those using PBO-LLINs had lower odds of malaria infection than those using a standard LLIN at both time points (t28; OR: 0.43, 95%CI (0.22-0.84), p value: 0.0151 and t33; OR: 0.34, 95%CI (0.16-0.71), p value: 0.0051). Similar findings were observed when comparing each of the PBO-LLIN arm individually to the standard LLIN arm (Table S3). There was no difference in malaria prevalence between the IRS arms vs no IRS arms at any of the time points in the ITT analysis (t28: 74.7% vs 76.0%, OR 0.79, 95%CI: 0.44-1.41, p value: 0.4172, t33: 51.6% vs 56.8%, OR: 0.52, 95%CI: 0.25-1.07, p value: 0.0747) or the PP analysis (Table 1). There was no significant reduction in anaemia prevalence in children under 5 years old in any of the intervention arms (PBO-LLIN or IRS arms) vs their respective comparators (Table S4). Post hoc intention to treat analysis comparing the PBO-LLIN groups to standard LLIN groups over time showed strong effect on malaria prevalence 9, 16, 21, 28 months post intervention, while at 4 months and 33 months there was no significant differences (Figure 2).

**Table 1:** Malaria infection prevalence in children 6 months to 14 years of aged at 28 and 33 months post intervention in intention to treat and per protocol analysis.

| Intervention | Intention to treat |  |  |  |  | Per protocol* |  |  |  |  |
| --- | --- | --- | --- | --- | --- | --- | --- | --- | --- | --- |
|  | n/N | % | OR | 95%CI | P-value | n/N | % | OR | 95%CI | P-value |
| **Survey 28 months** |  |  |  |  |  |  |  |  |  |  |
| Standard LLIN <sup>1</sup> | 1643/2032 | 80.9% | 1 |  |  | 762/962 | 79.2% | 1 |  |  |
| PBO LLIN <sup>2</sup> | 1280/1848 | 69.3% | 0.45 | 0.21-0.95 | 0.0364 | 507/755 | 67.2% | 0.43 | 0.22-0.84 | 0.0151 |
| No IRS <sup>3</sup> | 1441/1897 | 76.0% | 1 |  |  | 663/881 | 75.3% | 1 |  |  |
| IRS <sup>4</sup> | 1482/1983 | 74.7% | 0.79 | 0.44-1.41 | 0.4172 | 606/836 | 72.5% | 0.71 | 0.39-1.30 | 0.2615 |
| Interaction coefficient |  |  | 1.38 | 0.53-3.69 | 0.505 |  |  | 1.59 | 0.62-4.07 | 0.3239 |
| **Survey 33 months** |  |  |  |  |  |  |  |  |  |  |
| Standard LLIN <sup>1</sup> | 1045/1784 | 58.6% | 1 |  |  | 291/509 | 57.2% | 1 |  |  |
| PBO LLIN <sup>2</sup> | 901/1807 | 49.9% | 0.60 | 0.32-1.13 | 0.1131 | 200/453 | 44.2% | 0.34 | 0.16-0.71 | 0.0051 |
| No IRS <sup>3</sup> | 1009/1776 | 56.8% | 1 |  |  | 257/500 | 51.4% | 1 |  |  |
| IRS <sup>4</sup> | 937/1815 | 51.6% | 0.52 | 0.25-1.07 | 0.0747 | 234/462 | 50.7% | 0.57 | 0.31-1.06 | 0.0736 |
| Interaction coefficient |  |  | 1.78 | 0.71-4.49 | 0.2131 |  |  | 3.11 | 1.18-8.20 | 0.0230 |

OR, odds ratio for the factorial analysis compared the two-main intervention effect 1/PBO LLIN vs No PBO LLIN and 2/IRS vs no IRS and their interaction. OR unadjusted for baseline plasmodium infection prevalence.\* per protocol includes only children sleeping under the allocated nets.
<sup>1</sup> Standard LLIN and standard LLIN & IRS arms, <sup>2</sup> PBO LLIN and PBO LLIN & IRS arms, <sup>3</sup> standard LLIN and PBO LLIN arms, <sup>4</sup> standard LLIN & IRS and PBO LLIN & IRS arms.

**Table 2:** Entomological outcomes comparing the two main interventions (PBO LLIN vs No PBO LLIN arms and IRS vs No IRS arms) in 2017.

|  | Vector density per night per household |  |  |  |  | Sporozoite rate |  |  |  |  | EIR per nights per household |  |  |  |  |
| --- | --- | --- | --- | --- | --- | --- | --- | --- | --- | --- | --- | --- | --- | --- | --- |
|  | N | Mean | DR | 95%CI | P-value | n/N | % | OR | 95%CI | P-value | N | Mean<sup>5</sup> | DR<sup>6</sup> | 95%CI | P-value |
| Standard LLIN <sup>1</sup> | 1575 | 5.24 | 1 |  |  | 108/2070 | 5.2% | 1 |  |  | 1540 | 0.15 | 1 |  |  |
| PBO LLIN <sup>2</sup> | 1575 | 5.48 | 1.59 | 0.80-3.16 | 0.1832 | 67/1788 | 3.8% | 0.88 | 0.55-1.42 | 0.5992 | 1536 | 0.11 | 0.63 | 0.25-1.61 | 0.3296 |
| No IRS <sup>3</sup> | 1567 | 5.00 | 1 |  |  | 103/1962 | 5.3% | 1 |  |  | 1524 | 0.14 | 1 |  |  |
| IRS <sup>4</sup> | 1583 | 5.71 | 1.06 | 0.48-2.37 | 0.8807 | 72/1896 | 3.8% | 0.87 | 0.52-1.47 | 0.609 | 1552 | 0.12 | 0.97 | 0.41-2.28 | 0.9426 |
| Interaction coefficient |  |  | 0.47 | 0.16-1.40 | 0.1699 |  |  | 0.63 | 0.31-1.28 | 0.195 |  |  | 0.88 | 0.24-3.21 | 0.8489 |

Density ratio (DR) for vector density and EIR and odd ratio (OR) for sporozoite rate are adjusted for their respective baseline value. <sup>5,6</sup>Arithmetic mean and DR of the EIR are weighted to account the proportion of mosquitoes sampled to be tested for sporozoites. <sup>1</sup> Standard LLIN and standard LLIN & IRS arms, <sup>2</sup> PBO LLIN and PBO LLIN & IRS arms, <sup>3</sup> standard LLIN and PBO LLIN arms, <sup>4</sup> standard LLIN & IRS and PBO LLIN & IRS arms.

**Figure 2:**
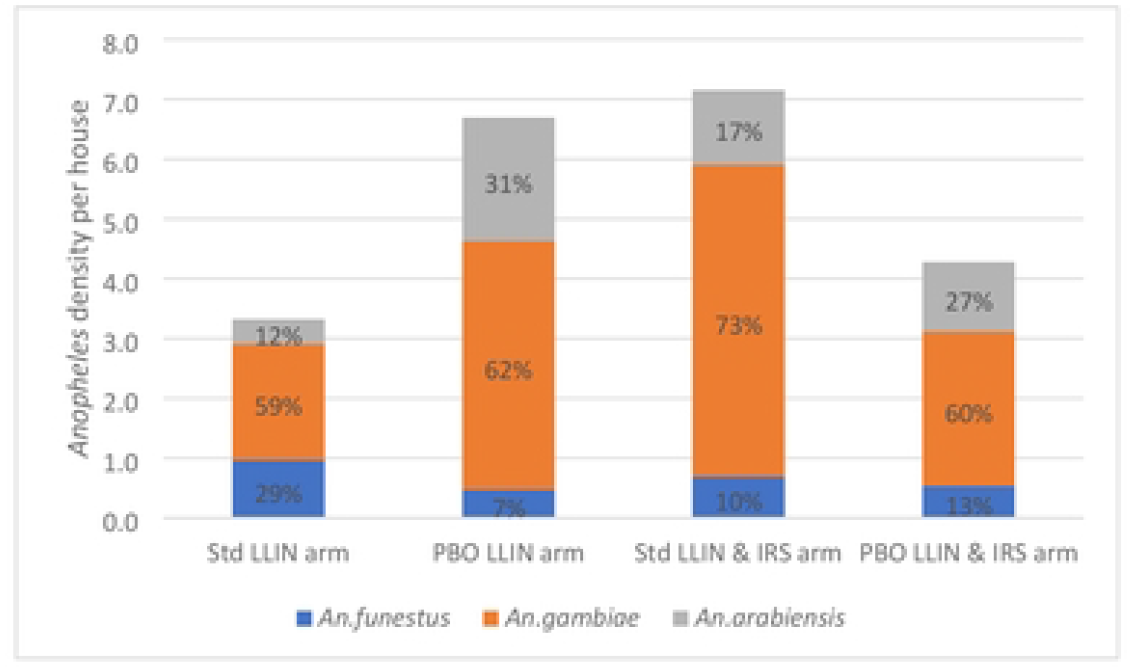
*Anopheles* densities per night per house for each vector species, proportion within arm also presented in each of the bar chart.

### Entomological results

A total of 31,120 mosquitoes were collected during the 3150 collection nights, of which 56% (N=17451) were identified as *Anopheles*. Vector densities were 3.3 per house per night in the standard LLIN arm, 6.7 in PBO LLIN arm, 7.2 in the arm with standard LLIN + IRS and 4.3 in PBO-LLIN + IRS arm (Figure 2). The proportion of mosquitoes positive for *falciparum* sporozoites was 2.6% for the combined arm PBO-LLIN + IRS and 4.9% and over for the other arms. In the factorial analysis there was no differences in vector density, sporozoite rate or EIR between PBO-LLIN vs standard LLIN arms or IRS vs no IRS arms during the third year follow up (Table 3). No significant effect was observed when comparing each of the individual arms to standard LLINs (Table S5). 87.6% (14784/16884) of female vectors were *Anopheles gambiae* sensu lato and the remaining (2100/16884) were *An. funestus* s.l.. Of the 892 *An. gambiae* s.l. tested by PCR to distinguish between sibling species, 75% were *An. gambiae* s.s. (N=670) across the 4 arms whilst the highest proportion of *An. arabiensis* was found in the two arms with PBO-LLINs.

### Harms and limitations

Limitations, the number of children in the PP analysis is half of those in the ITT analysis at t28 and only a quarter at t33.

## Discussion

In this CRT assessing the effectiveness of a new class of LLIN over 3 years, children residing in clusters that received PBO-LLINs had reduced odds of malaria infection compared to those who received standard LLINs up to 28 months post-intervention when half of the participants were still using the study nets. This effect was however weaker and no longer significant 6 months later when usage has dropped further and 96% of the PBO concentration was lost. However, odds of infection in children still sleeping under the PBO-LLINs remained significantly lower compared to those sleeping under standard LLIN over this third year. There was no reduction in vector density and malaria transmission (estimated through EIR) in the PBO-LLIN vs standard LLIN groups. This contrasted with the first two years of the CRT where reductions were observed in both malaria prevalence, and malaria transmission in the PBO-LLIN groups vs the control groups [7], suggesting a large effect on vector population density and protection extended to the overall community when PBO-LLIN were less than 2 years old and usage over 40%. A malaria transmission model showed that net usage as low as 35% can provide a mass effect [27] and those have also been reported at high net coverage in field studies [28, 29].

In our study, as net usage dropped in the third year and concentrations of PBO and permethrin declined, this community effect might have been lost and only individual protection remained, with the net providing a physical barrier against mosquito bites. This may explain the lack of impact of PBO-LLINs on malaria transmission and the moderate protection observed mainly in children using PBO-LLINs. Permethrin content in the PBO-LLINs and standard LLINs were halved after 36 months, while PBO was reduced by more than 95% of the initial concentration. These insecticidal contents were comparable to those reported after 36 months of use in a study conducted in Kenya [19] with the same brand of net. The faster reduction of PBO compared to pyrethroid was also observed in another community trial in Uganda [32] and similarly reported when nets were washed in laboratory conditions [33-35]. This raises concerns about the long-term chemical durability of PBO and how long this new class of net will provide a superior effect compared to standard LLINs in areas of pyrethroid resistance. The Ugandan study was part of a CRT and showed higher mortality in resistant *Anopheles* exposed to Olyset Plus vs Olyset net up to 25 months when the PBO concentration was around 4 g/kg [32], which was associated with significantly lower malaria prevalence and vector density [8]. The authors also observed a direct correlation between PBO content and *Anopheles* mortality with reductions in mosquito killing effect by 11% for each gram per kilogram of PBO lost [32]. In the first two years of our trial, an effect of PBO-LLIN on malaria transmission and vector density was observed when concentrations of PBO were reduced to 16% of its original content. Personal protection against *Anopheles* bites was still observed in torn PBO-LLIN used for 21 months in the field [36] this may be due by higher excito-repellency offered by PBO-LLIN compared to standard LLIN [37]. It has been suggested that higher pyrethroid release rate in this specific brand of PBO-LLIN (Olyset Plus) may increase the concentration of the insecticide on the surface of the net fibre and therefore have a higher mosquito killing effect compared to Olyset net, regardless of PBO concentration [34]. Pyrethroid contents remaining in nets used for 24 months was much lower in Olyset Plus (6 g/kg) vs Olyset net (16 g/kg), and if bioavailable on the net surface instead of contained within fibres, may potentially explained the superior personal protection of Olyset Plus in the third year. However, at the end of 36 months, permethrin contents were comparable between LLINs (9 g/kg and 10 g/kg in Olyset Plus vs Olyset net, respectively), suggesting protection provided to PBO-LLIN users may instead be attributable to the barrier effect (if remaining nets were still in good condition). Alternatively, the pyrethroid bleed rate in PBO-LLINs may have slowed after 36 months or our sample size of 10 nets was not sufficiently representative of all ageing PBO-LLINs.

The reduction in effective lifespan of PBO-LLINs and standard LLINs was also observed in a cohort of nets followed annually as part of this study showing less than 30% of the nets distributed survived up to 3 years as expected by WHO [25]. It also showed that the development of net holes was not associated with increased exposure to malaria in our study area, but was the main driver of decreasing usage [25]. Similarly, others did not observe a direct association between net textile deterioration and malaria [38, 39], while level of net attrition was related to the accumulation of net wear and tear [18]. A modelling study concluded that low net usage may have the greatest impact on net effectiveness against malaria vectors, rather than holes or bio-efficacy [40]. These findings are corroborated by our most recent study conducted in Tanzania where PBO-LLINs effectiveness was only observed for a year, as net usage drastically dropped in subsequent year [12].

The WHO currently requires CRTs to be conducted over a minimum of two transmissions seasons to assess the public health value on a new class of LLIN. However, NMCPs must collect more information about net efficacy against malaria vectors in the third year of intervention use, as well as conduct routine assessments of the impact of aged nets in the field; unless the procurement and distribution regimens change from 3 years to a more accurate effective lifespan of a product. The Tanzanian continuous delivery mechanism of LLINs through yearly school distributions to primary children and during antenatal consultations, rather than mass distribution seems more adapted to the PBO-LLIN lifespan by maintaining good coverage and a mixture of newer effective nets and aged net across the community [41]. The development of stronger net material would also improve net usage if holes were to appear more slowly and, finally, community sensitisation to improve care and repair could also reduce the rate of textile degradation [42, 43], and maintain higher coverage, thereby increasing the length of time between distributions. Intervening in each of these parameters will alter the relative cost-effectiveness of products and the viability of each strategy [44].

The absence of effect observed in the IRS arm was expected as pirimiphos-methyl does not last on house walls more than 9 months [45] and yearly rounds of IRS are usually necessary to maintain efficacy, but have cost implications. A modelling study, based on the present CRT area characteristics and malaria prevalence, reported that the effect of repeated IRS would have averted 45 more malaria cases per 1000 people per year over 3 years, compared to PBO-LLINs alone, and this benefit would have been the highest in the third year, when net usage had decreased [44]. However, PBO-LLINs were still reported as being the most cost-effective intervention. Longer-lasting formulations of non-pyrethroid insecticide for IRS might be a more cost-effective alternative to existing IRS products. New IRS active ingredients (for example, clothianidin and broflanilide) also have potential to be used in resistance management schemes with PBO-LLINs, if no antagonistic effects are present [46]. Deployment regimens for IRS and new dual-active-ingredient LLINs need to more accurately reflect the true effective lifespan of each product to avoid leaving both LLINs and IRS in communities past their optimum operational lifespans. Without timely replacement, vector populations may be exposed to sublethal insecticide doses with the potential for faster selection of insecticide resistance [47].

PBO-LLINs still provide personal protection to user by the third year but did not reduce malaria transmission compared to pyrethroid-only LLINs and malaria prevalence was unacceptably high. Campaigns based on LLIN replacement every three years are unlikely to control malaria adequately and will leave populations unprotected. For maximum impact, distribution regimens must be adapted to the effective lifespan of new classes of LLINs products coming into the market and / or longevity of this product should be prolonged to meet the WHO recommended chemical and textile durability of three years.

## Data Availability

Data cannot be shared publicly because of requirement for data transfer agreement. Data supporting this research may be provided upon request to the primary author and completion of data transfer agreement from the Tanzania National Institute of Medical Research (http://reims.nimr.or.tz:8010/guides/DTA.pdf).

## Contributors

NP, IK, MR conceived and designed the study. FWM, CDM, WK advised on interventions, study communities and coordination with local and national authorities. NP, JFM, EL, JDC, EK, AW, AM implemented the study. NP and JFM analysed the data. NP, JFM, IK, MR, LAM, EK interpreted the data. NP, JFM & LAM wrote the first draft of the manuscript. MR & IK critically revised the manuscript for important content. EL, DJC, AW, CDM, AM, FWM, EK, WK revised the manuscript. All authors read and approved the final version of the manuscript.

## Acknowledgements

The authors express their sincere thanks to colleagues of Kilimanjaro Christian Medical University College and National Institute of Medical Research who were involved in the project for their hard work. We acknowledge the support provided by governmental authorities in Muleba District. We thank the study trial steering committee members. Thank you to Jackie Cook (LSHTM) and Manisha Kulkarni (U-Ottawa) for their advice on some analysis component. We wish to thank all the participants. This study was funded under the Joint Global Health Trial scheme by Foreign, Commonwealth and Development Office (FCDO), the Medical Research Council and Wellcome. (Grant Ref: MR/L004437/).

---

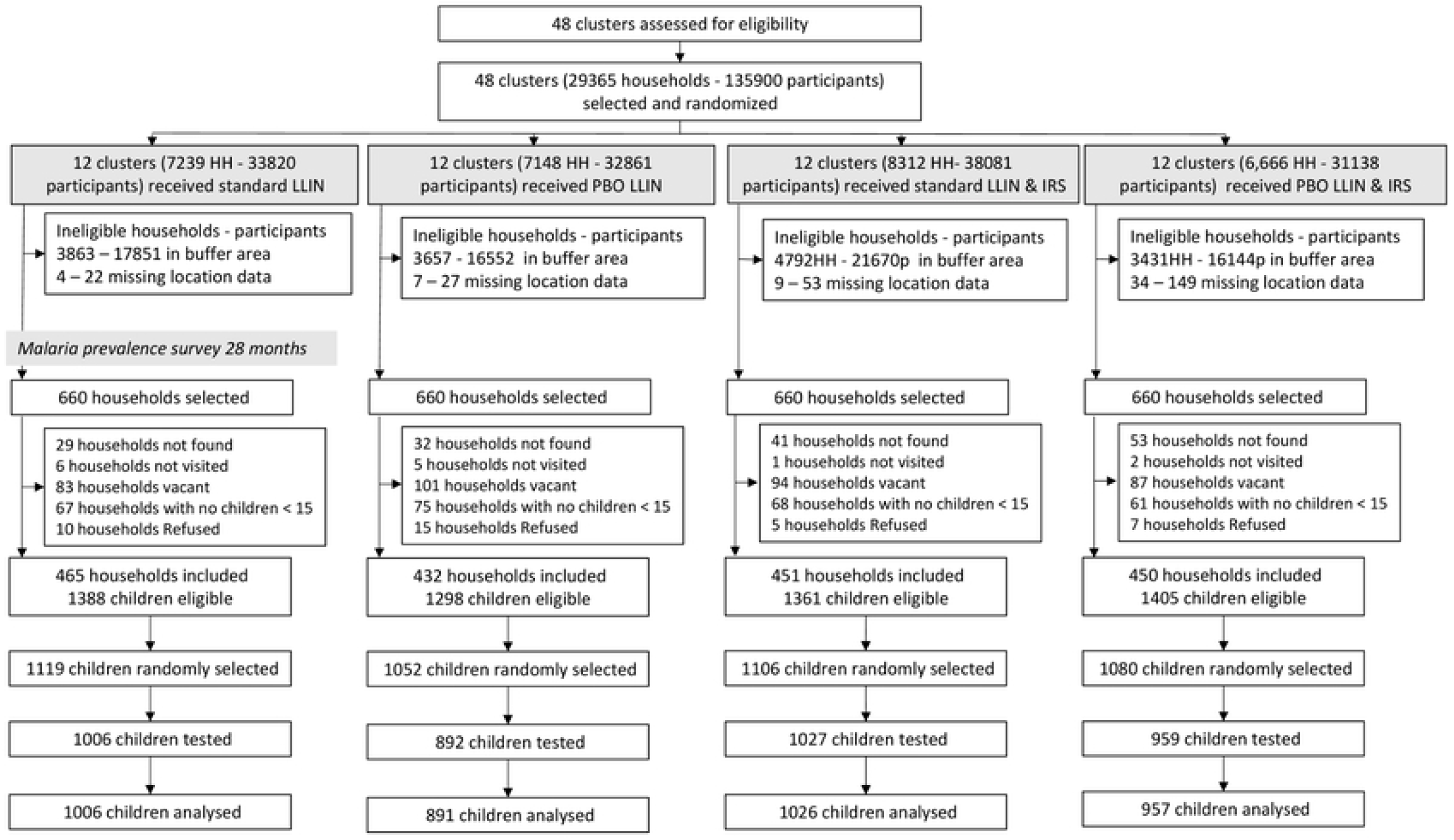

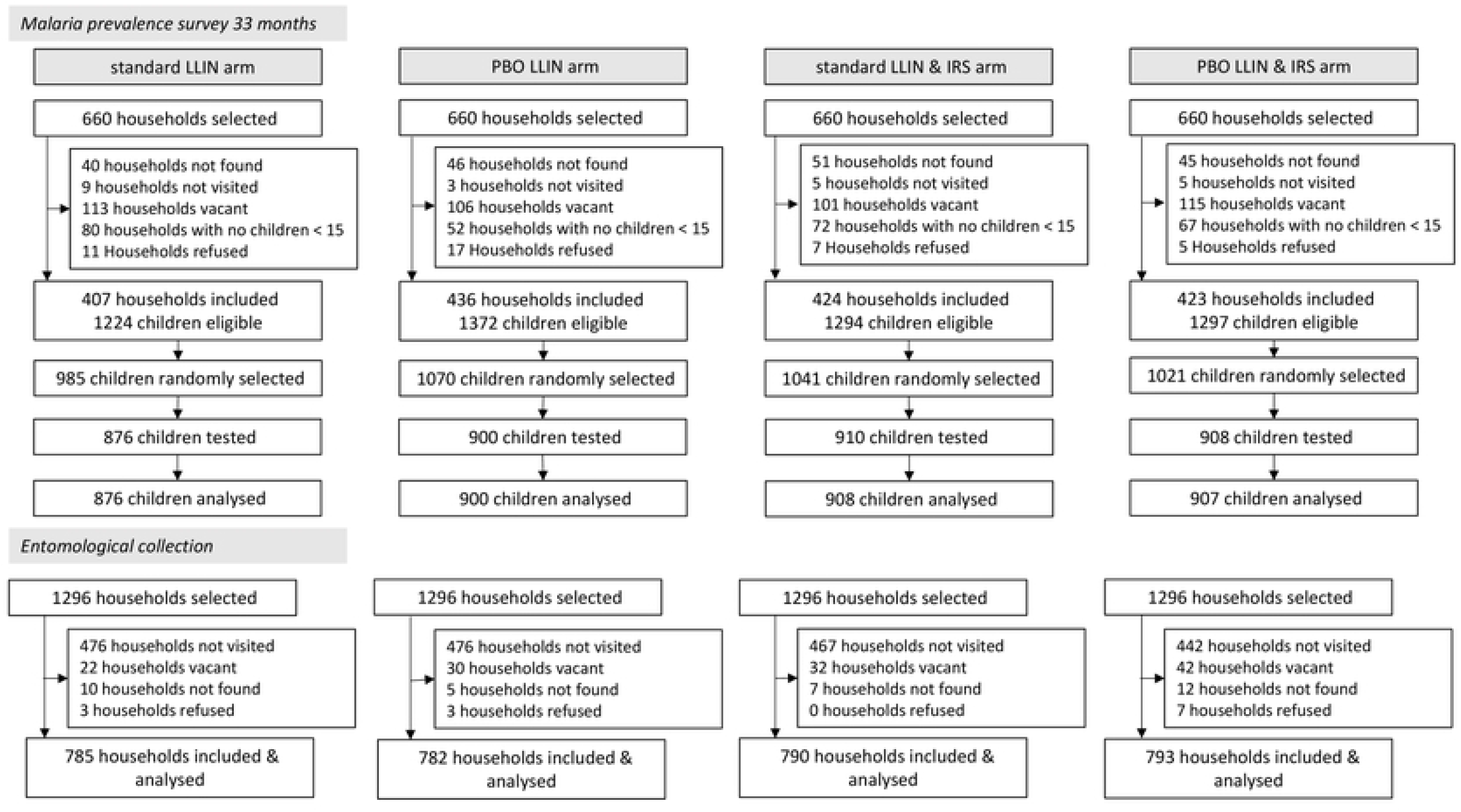

